# Activation Metrics for Structural Connectivity Recruitment in Deep Brain Stimulation

**DOI:** 10.1101/2025.01.16.25320420

**Authors:** Konstantin Butenko, Jan Roediger, Bassam Al-Fatly, Ningfei Li, Till A. Dembek, Gan Yifei, Zhu Guan-Yu, Zhang Jianguo, Andrea A. Kühn, Andreas Horn

## Abstract

Comparatively high excitability of myelinated fibers suggests that they represent a major mediator of deep brain stimulation effects. Such effects can be modeled using different levels of abstraction, ranging from simple electric field estimates to complex multicompartment axon models. In this study, we explore three approaches to estimate axonal activation: electric field magnitudes, electric field projections and (probabilistic) pathway activation modeling. Our aim is to describe these approaches and also illustrate their relevance. For that, we apply them to a clinical dataset of 15 Parkinson’s disease patients, who were stimulated in the subthalamic nucleus in bipolar mode. To make these approaches accessible for the community, necessary modeling and statistical processing was implemented in the openly available Lead-DBS toolbox.

## Introduction

In deep brain stimulation (DBS), various modeling approaches have been considered to quantify effects on neural tissue, and, in particular, on white matter tracts. Based on the Hodgkin-Huxley model ^1^, an activating function was proposed defined by the second spatial derivative of the extracellular electric potential along the fiber ^2^. Furthermore, to capture an accumulated effect of the stimulation along the fiber, driving force methods were proposed ^3,4^, which operate with weighted sums of the second finite differences of the potential. For a monopolar single contact case, however, the activation may also be approximated by the electric field magnitude ^5^. Increasingly, studies have used binarized versions of the magnitude (often at 200 V/m ^6,7^) to delineate an all-or-nothing stimulation volume. Mistakenly, this thresholded version of the electric field magnitude has sometimes been referred to as the volume of tissue activated (VTA), which, strictly speaking, is a different concept proposed earlier in ^8^. In its original definition, the VTA quantified activation extents based on a stimulus response of axon models that were placed on a regular grid around the electrode. In a more advanced concept of pathway activation modeling (PAM), such axons are not placed on a grid but follow anatomically meaningful trajectories of white matter tracts ^9^. Of note, such white matter streamlines could be defined based on diffusion-weighted imaging (DWI) ^10^ or, instead, based on manually curated and more accurate anatomical pathway atlases ^11^.

To make an appropriate choice from this large repertoire of available methods, an investigator has to understand their relevance and limitations for the respective goals and hypotheses of a given study ^12^. Indeed, goals of study may starkly differ, from estimating DBS outcome over gaining anatomical insights into brain regions and networks involved in therapeutic neuromodulation to understanding detailed biophysical effects on differentiated pathways. Hence, there may not be a clear one-size-fits-all solution to the problem. Beyond their differing complexity and biophysical plausibility, a key additional component to consider is whether the respective model results in a binary (‘all or nothing’) or continuous (probabilistic) estimate of tract activations. Either type of result will dictate statistical tests for a group-level analysis such as probabilistic mapping ^13^, DBS network modeling ^14^ or fiber filtering ^15,16^.

Modeling activations in an all-or-nothing fashion may have the advantage of strict compatibility with biology in the sense that axons fire in an all-or-nothing fashion. However, one downside may be that these binary decisions require a list of assumptions such as the fiber diameter, tissue conductivity and dispersivity, electrode-tissue interface, just to name a few. Hence, while the response may seem clean and biophysically plausible, a downside is that the model might fail to capture reality if even a single assumption is ill-informed. Critically, streamlines in computer simulations could also be conceptualized as axonal *populations*, of which certain subpopulations – such as the ones with thicker myelination – may be activated at lower stimulation amplitudes than the rest of the population. This view would render probabilistic forms of modeling compatible with biology and may bear advantageous possibilities for the downstream statistical analyses ^17^. Indeed, each fiber bundle in a standard neuroimaging analysis represents 10^3^−10^5^ tightly packed axons ^18^ with varying fiber diameters ^19,20^. And even if diameters of some neuron populations may fall within a narrow range, it may still be of utility to model activations in probabilistic fashion to keep models robust against ill-informed or simplistic assumptions, such as exact axonal diameters, lengths, etc. Finally, a practical disadvantage of *binary* (‘all-or-nothing’) activation models for group statistics is that they require splitting the patient cohort into two subgroups in which the fiber was either activated or not ^21^. Unfortunately, this split of patients into two groups happens differently for each bundle, making resulting statistical tests, e.g., two-sample t-tests, less comparable across fibers. This problem is even more accentuated for small cohorts, in which two-sample tests would compare even smaller subcohorts, or for fibers that split the cohort into lopsided subcohorts (e.g., comparing 95% of ‘non-activated’ vs 5% of ‘activated’ patients in the cohort). This practical disadvantage requires setting heuristical parameter choices, such as a minimum or maximum amount of activations of the tract across the groups, which may lead to both type I and II errors when associating tracts with clinical improvements. Instead, continuous activation models have the practical advantage as they allow regression or correlation to be conducted for the entire patient cohort when analyzing relationship between probabilities of activations and clinical scores ^17^.

In this paper, we describe three probabilistic activation metrics and showcase them in the context of a group level analysis. These metrics are the “classic” electric field magnitude, the electric field projection, which quantifies the depolarization gradient along the fiber, and an adaptation of pathway activation modeling, which incorporates uncertainty of axon model parameters (probabilistic PAM, or pPAM). As all three metrics result in continuous (probabilistic) estimates of pathway activations, they can be directly compared to one another since they allow application of the exact same group-level statistics. For sake of completeness, binarized activation models (thresholded electric fields and binary PAM) were also evaluated. To illustrate the metrics and discuss their applicability, we conduct their comparison using clinical data of Parkinson’s disease patients implanted in the subthalamic nucleus and stimulated using bipolar mode. Bipolar settings were deliberately chosen as they represent a more complex modeling case ^22^. The analysis reveals a discrepancy in the identified structural connectivity and its association with clinical outcomes. To facilitate future studies, these metrics are implemented in the open-source toolbox Lead-DBS ^23^.

## Materials and Methods

### Concepts

DBS simulation in a volume conductor model provides a distribution of the extracellular electric potential ^24^, which can be used to derive an electric field vector, and, subsequently, the electric field magnitude || E||, which has been explored as a direct proxy for axonal activations, in the past ^5,17,22^. However, activation of a fiber is induced by the electric field component *parallel* to it ^25,26^, so projecting the field onto the fiber is a potentially more informative metric of the DBS effect. This can be computed as the dot product of the electric vector field with the unit vector tangential to the fiber (see *Fig. 1*). Note that both the magnitude and the projection are evaluated at multiple points along the fiber, and a fiber-wise value can be defined as either a sum, mean or peak of these point-wise field metrics. This fiber-wise value may then be used as a (continuous) regressor in a group level analysis, which aims at setting DBS effects into relation with empirically observed stimulation outcomes ^15,16^.

**Figure 1:**
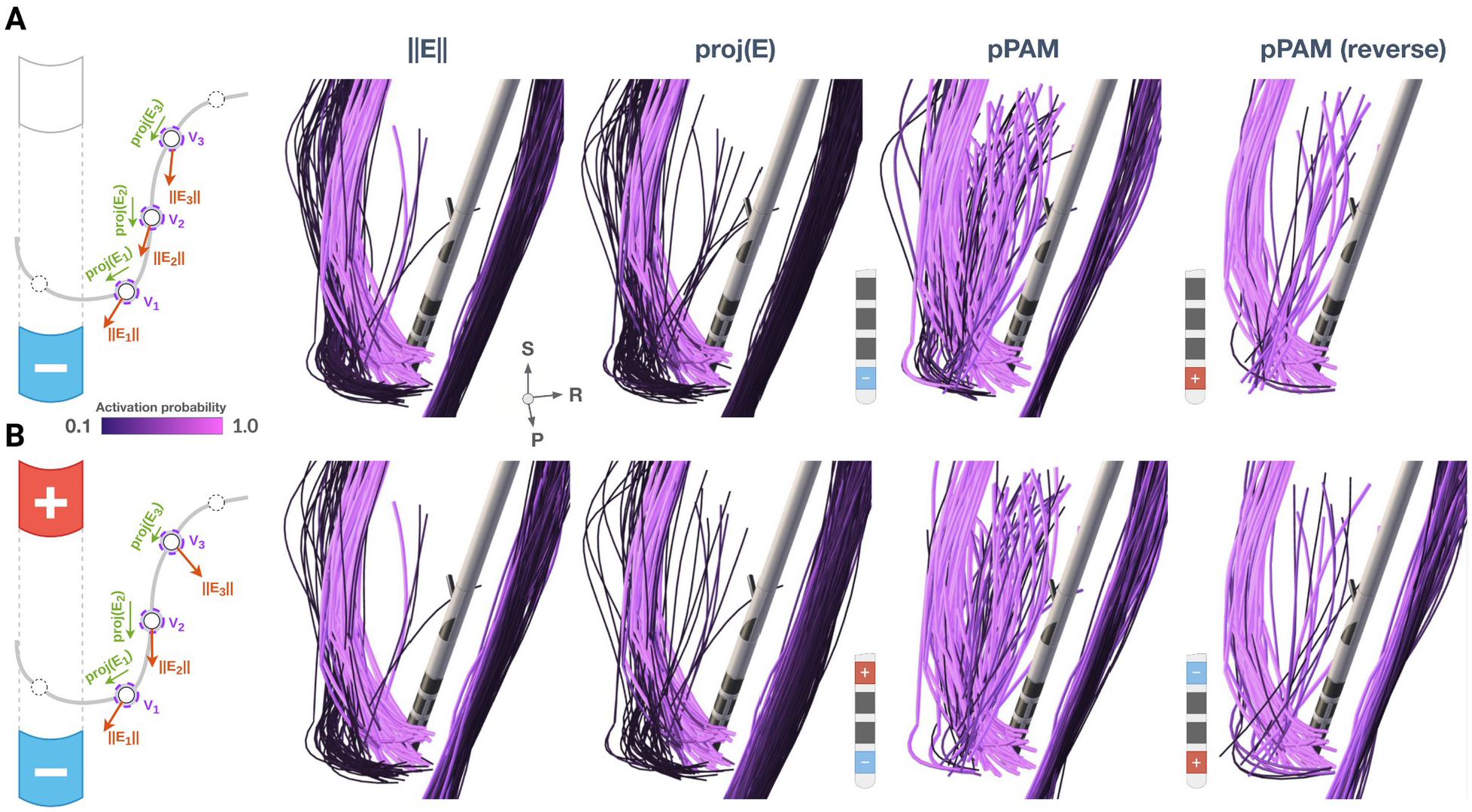
Differences between the three activation metrics for monopolar (**A**) and bipolar stimulations (**B**). **Left:** schematics of the activation metrics. The electric field magnitude depends on the stimulation settings and the volume conductor model, while the electric field projection proj(E) also accounts for the local directionality of the fiber in relation to the directionality of the field. The directionality is also encoded in the axonal activation model, which computes the change in the membrane potential in response to the stimulus-induced distribution of the extracellular potential along the fiber. Axonal activation models rely on multiple parameters and assumptions, e.g., myelination thickness, spacing between nodes of Ranvier, ion channel dynamics, etc. Instead of selecting particular values, these parameters can be sampled probabilistically, thus providing a probabilistic estimation of activation (pPAM). **Right:** Distribution of the activation metrics across selected hyperdirect, pallidosubthalamic and corticospinal fibers sampled from the Basal Ganglia Pathway Atlas ^11^. While the distribution is generally similar for the magnitude and projection metrics, the difference can be observed for the pallidosubthalamic fibers, especially in the bipolar case. Anodic stimulation (estimated by pPAM) leads to lower activation of fibers passing or parallel to the lead (panel A, right). For bipolar stimulation, lower activation is predicted for short passing fibers when they terminate closer to the anode (panel B, right). Critically, the electric field based metrics cannot account for polarity.

However, neither the magnitude of the electric field ||E|| nor its projection onto the fiber *proj*(E) may encode simultaneous depolarization and hyperpolarization effects introduced by bipolar stimulations. For this, a more complex concept of pathway activation modeling has to be used. First, PAM uses a time domain solution of the electric field problem to address the dispersion (frequency dependency) of the brain tissue conductivity ^27^. This can be achieved by using the *Fourier Finite Element method* that computes solutions for multiple frequencies that compose the DBS signal power spectrum ^8^, which can be, however, truncated ^28^. Second, beyond modeling the electric field, a computational axon model has to be used, which, in turn, requires specification of multiple parameters, such as ion channel characteristics, myelin dielectric properties, internodal distances, etc. These parameters are subject of uncertainties, and, therefore, we propose to model PAM in a probabilistic fashion. For example, fiber diameters may be sampled from a normal or uniform distribution of values reported in the neuroanatomical literature. Once deterministic parameters have been specified and probabilistic parameters have been sampled, the cable equation of the axon model can be solved in order to obtain an estimated change in the membrane potential in response to the alteration of the extracellular potential along the fiber. If an action potential is detected, the axon is considered “activated”. Note that solving such cable equations using finite difference methods is (computationally) relatively cheap and easily parallelizable on modern multicore machines making a probabilistic assessment with different parameters feasible even for larger cohorts.

Probabilistic pathway activation modeling may inform the certainty of the axonal activation for the given probabilistic parameters. The certainty can be simply estimated as *N*_activated_ / *N*_samples_, where *N*_activated_ - number of samples, in which the axon is deemed activated, i.e., responded with an action potential. For the simpler electric field metrics, field strengths may be converted to activation probabilities using a sigmoidal function ^29^: the higher the fiber-wise electric field value the higher the certainty of its activation. Although this approach does not explicitly model probabilistic axonal parameters, it accounts for the fact that different axonal morphologies have different activation thresholds for extracellular stimulation ^5^. The determined certainty of activation may be leveraged in a statistical analysis, e.g., by using it as a weight in a linear regression for continuous variables or in odds ratio analysis for binary variables ^29^.

### Notes on Implementation

The described methodology was implemented in the Lead-DBS pipeline with the following steps. First, non-linear warps from the patient (native) to standard space were obtained and electrodes were localized following the Lead-DBS pipeline ^23^. Second, a structural connectivity atlas ^11^ was non-linearly warped into patient space. Then, electric field metrics and pathway activations were computed in patient space using OSS-DBS software ^24^. The latter was conducted *N* times for *N* probabilistic samples. To probe action potentials in response to the stimulation, a NEURON ^30^ implementation of McNeal’s mammalian axon model was employed ^31^.

For the statistical group-level analysis, the results were imported into the Fiber Filtering Explorer ^23^. This tool operates in the normative MNI space, but since the activation metrics were defined on the fibers (from the warped structural connectivity atlas), native space results could be imported directly. This is a new feature of the toolbox, while previous fiber filtering studies in Lead-DBS have evaluated activations of fibers based on their intersection with the electric field in template space. The fiber-wise metrics may also be “mirrored” across hemispheres, if the structural connectivity atlas itself is available in streamline-mirrored fashion, i.e., each streamline from one hemisphere has a direct counterpart in the other hemisphere.

### Clinical Data

To explore the structural connectivity metrics on a group level, we retrospectively analyzed 15 Parkinson’s patients who had undergone bilateral STN-DBS electrodes implantation at Charité – Universitätsmedizin Berlin and Beijing Tiantan Hospital. Patients were selected based on the availability of imaging data and clinical motor assessment, and only patients with bipolar stimulation settings were included. The collection and analysis of all patient data was approved by the Local Ethics committee of Charité – Universitätsmedizin Berlin (master vote EA2/186/18) and the Local Ethics Committee of Beijing Tiantan hospital (KY2022-006-02). All patients were implanted with omnidirectional four level electrodes: 14 – Medtronic (Minneapolis, MN, USA), 1 – PINS (Beijing, China). For details on demographics, clinical outcomes and stimulation protocols, please, refer to *Suppl. Table 1*. Based on the pre- and post-operative MRI and post-operative CT imaging, DBS electrodes were localized following the standard pipeline implemented in Lead-DBS ^23^. This included computation of non-linear warps between the patient (native) space and template (MNI) space using Advanced Normalization Tools ^32^, a brain shift correction to account for possible pneumocephalus and, if necessary, a manual correction of the warp field using the WarpDrive tool ^33^. To describe the structural connectivity around the STN region, the Basal Ganglia Pathway Atlas was employed ^1^. Electric fields and pathway activations were computed in a heterogeneous and dispersive volume conductor model using OSS-DBS software ^24^ (https://github.com/SFB-ELAINE/OSS-DBSv2.git), and results were further processed as described in the previous section. To also model and compare *binary* activation metrics, electric field magnitudes were thresholded at the commonly used 200 V/m ^5^, while the threshold for electric field projections was set to 125 V/m in order to match the total number of fibers recruited by these two metrics in at least 20% of patients. Probabilistic PAM was conducted for uniformly sampled fiber diameters (N = 10) in the range of 1–4 um defined based on the neuroanatomical literature ^19,20,34^. For binary PAM, two thresholding levels were used: with p(A) = 1.0 (the axon was activated for all fiber diameters) and p(A) ≥ 0.1 (the axon was activated for at least one fiber diameter). The clinical effect of the structural connectivity recruitment was described by the percent improvement of Unified Parkinson’s Disease Rating Scale III score (motor examination, UPDRS-III) between DBS-on and DBS-off conditions, both evaluated in the medication-off state. For each fiber, an unpaired two sample t-test and its probability-weighted version were applied to compare outcomes across patients, in which the fiber was “activated” and the ones in which it was not. Only fibers “activated” (for probabilistic metrics p(A) ≥ 0.5) in at least 20% of stimulations were considered to avoid model overfitting. To quantify differences and similarities of pathway activation profiles, i.e., percent activations across pathways, paired sample t-tests and Spearman’s rank correlations were used.

## Results

Group-level results are shown in *Fig. 2*. All electrodes were localized in the subthalamic region, primarily in the dorsolateral aspect of the nucleus (*Fig. 2 panel A*). Using each metric (||E||, *proj*(E) and PAM), profiles of percent activations of all pathways represented in the atlas were computed (*Fig. S1)*. These profiles demonstrated varying degrees of agreement across patients and hemispheres. *Figure 2, panel B* shows rank correlations between profiles assessed at ||E|| ≥ 200 V/m and PAM, with only 21/30 instances being significantly correlated (uncorrected p < 0.05). Particular profiles for two selected patients are shown in *Fig. 2, panel C. Proj*(E) and PAM showed a similar agreement (20/30, see *Fig. S2*), and the paired sample t-test on the z-scored rank correlations showed non-significant difference between ||E|| and *proj*(E) correlations to the PAM profiles (p = 0.28). The ||E|| and *proj*(E) profiles were highly correlated (28/30), see *Fig. S3*.

**Figure 2:**
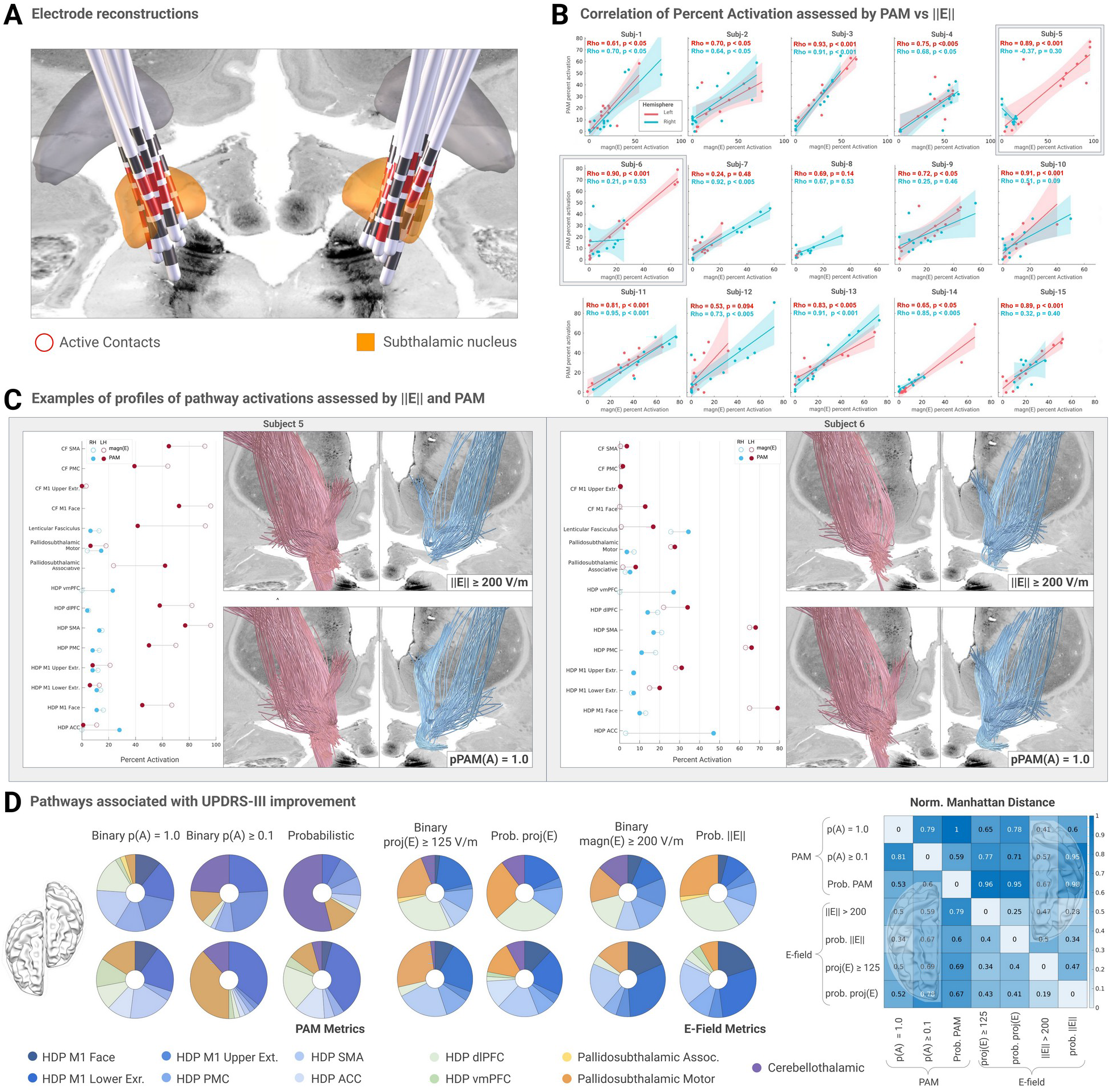
Group-level analysis of 15 Parkinson’s patients, who underwent STN-DBS. **A:** Using Lead-DBS, electrode trajectories were reconstructed showing localization to the subthalamic region. **B:** Correlations of pathway activation profiles computed for the applied bipolar stimulation protocols based on thresholded electric field magnitudes (≥ 200 V/m) and binary pathway activation modeling. Each dot denotes computed percent activation for one pathway using both metrics. While PAM and ||E|| approaches showed comparable results in most cases, for 9 out of 30 electrodes Spearman’s correlation of pathway activation profiles did not reach statistical significance and even showed an inverse correlations in one case (Subj-5) highlighted in **C:** Detailed description of the activation profiles computed using the electric field magnitude and PAM for example subjects 5 and 6. **D:** Donut (ratio) charts of pathways associated with UPDRS-III improvement across the activation metrics and statistical models. Ratios were computed based on 100 fibers (50 per hemisphere) with the highest T-values. To evaluate similarity across these models, Manhattan distances were computed, shown separately for each hemisphere. CF - corticofugal pathways, HDP - hyperdirect pathways, M1 - primary motor cortex, PMC - premotor cortex, SMA - supplementary motor area, ACC - anterior cingulate cortex, dlPFC/vmPFC - dorsolateral and ventromedial prefrontal cortex.

Statistical analyses of the pathways associated with UPDRS-III improvement also revealed clear differences across the metrics, see *Fig. 2, panel D*. The donut charts show pathways that contain 100 fibers (50 per hemisphere) with the highest T-values. In contrast to binary PAM, which assumes small fiber diameters (p(A) = 1.0), binary PAM for large diameters (p(A) ≥ 0.1) and the probability-weighted PAM emphasized the cerebellothalamic pathway, which is located posterior and dorsal to the subthalamic nucleus. Both binary and probabilistic *proj*(E) also identified this pathway, which was not always the case for ||E||. All metrics identified various branches of the hyperdirect pathway, converging on the motor projections, as well as the motor portion of the pallidosubthalamic tract. The heat map in

*Fig. 2, panel D* shows normalized Manhattan distances between these improvement associated pathway profiles. Note that the electric field metrics yielded more similar profiles, while the PAM metrics were in larger disagreement, especially when assuming small fiber diameters (p(A) = 1.0).

## Discussion

We must emphasize that our study was not designed to identify neural substrates underlying clinical effects of stimulation, since the amount of analyzed clinical data points (15 in total) would not yield robust statistical models that can be interpreted in neurophysiological and clinical context. Instead, we aimed at i) introducing an open-source implementation of discussed activation metrics in the commonly used DBS analysis pipeline ii) highlighting differences across activation metrics of structural connectivity in theoretical and also clinically relevant stimulation analyses.

The abstract example (*Fig. 1*) shows a prominent difference in pathway activations depending on the electric field orientation. The directionality is entirely ignored by the magnitude metric ||E||, but accounted for by the projection metric *proj*(E) and PAM. It should be, however, noted that the difference is less evident for monopolar stimulations, where the electric field configuration is more trivial. Another case is anodic stimulation that leads to a comparatively lower activation of passing ^35^ and parallel fibers (*Fig. 1A, right panel*). When the polarity was flipped in the bipolar mode so that the anode was closer to the tracts (*Fig. 1B, right panel*), the activation levels remained similar for fibers traversing in the immediate vicinity of the lead and for longer fibers parallel to the lead, while activation of the shorter passing fibers was decreased. These modeling results match clinical observations that higher anodic currents are typically required to achieve comparable clinical effects and side-effects as in cathodic stimulation ^36,37^. Importantly, the change of polarity would not affect predictions made by the electric field metrics, which has already been recognized as their major limitation ^22^.

To assess relevance of these theoretical considerations, we applied the discussed metrics to 15 STN-DBS patients, who were stimulated in bipolar mode. In general, electric field metrics ||E|| and *proj*(E) were in good agreement (*Fig. 2, panel D* and *Fig. S3)*. These results suggest that no selective pathway stimulation was achieved due to the field directionality alone. However, it should be noted that all patients had omnidirectional electrodes, so current-steering capabilities were limited. On the other hand, PAM yielded distinctly different activation profiles, and even an inverse correlation was observed in one instance (subject 5, right electrode). Furthermore, results of probabilistic PAM indicated a strong effect of the fiber diameter choice (see *Fig. 2, panel D*), which urges further investigation of the axonal morphology and modeling methodology for target and side-effect pathways. Lesser, though still prominent discrepancies were observed for the electric field metrics when comparing binary with probabilistic approaches (also related to the fiber diameter ^5^). It is worth mentioning, however, that all three metrics associated the motor hyperdirect pathways with motor improvement.

While PAM represents a biophysically more plausible approximation of the structural connectivity recruitment, its advantages for clinical applications have not yet been demonstrated. Furthermore, PAM requires significantly larger computational resources. The necessity to operate with time domain solutions limits applicability of computations on denser structural connectomes that may feature millions of streamlines. In contrast, electric field metrics can be interpolated from the solution probed at a highly dense grid of points, and only three values (vector components) per point are required. On the other hand, PAM uses just one value per spatial point, i.e., the scalar extracellular potential, but assessed at multiple time points (depending on the DBS signal resolution). This obstructs storing the solution on such highly dense grids (or degrees of freedoms of the finite element model), thus necessitating an a priori choice of specific fiber trajectories along which the solution is probed.

This work does not propose entirely new activation metrics but builds upon existing ones. Within the scope of this work, however, all metrics have been implemented into the same software framework and are now openly available to the community, which may facilitate research of novel stimulation paradigms. Of note, all three metrics are computed in patient (native) space to increase modeling accuracy. Moreover, they allow *probabilistic* estimations of activation that can be leveraged in the statistical framework of Lead-DBS. Finally, the described electric field metrics are not limited to DBS and could easily be extended for the use in other electric stimulation modalities.

## Data Availability

The code necessary to reproduce the findings of the study is available at https://github.com/netstim/leaddbs.git
The raw data are not publicly available due to patient privacy restrictions, but can be requested from the authors within the scope of a data sharing agreement.

## Conclusion

We illustrated and applied three activation metrics for structural connectivity analysis in deep brain stimulation. For activation assessment, theoretically, the projection of the electric field onto the fiber is more informative than the electric field magnitude. Practically, however, these metrics identified comparable structural connectivity profiles in the analyzed dataset of 15 STN-DBS patients stimulated in bipolar mode via omnidirectional electrodes. In contrast, pathway activation modeling resulted in distinctly different activation patterns, which, nevertheless, also associated stimulation of the motor hyperdirect pathway with UPDRS-III improvement. Furthermore, by implementing a probabilistic statistical framework into our analysis, we were able to assess a group-level effect of uncertainty in fiber diameters. This effect appeared to be prominent, which warrants further morphological studies of the neural substrate that is directly affected by deep brain stimulation.

