## Supplementary figures and images for "Activation Metrics for Structural Connectivity Recruitment in Deep Brain Stimulation"

### Supplementary Figure S1

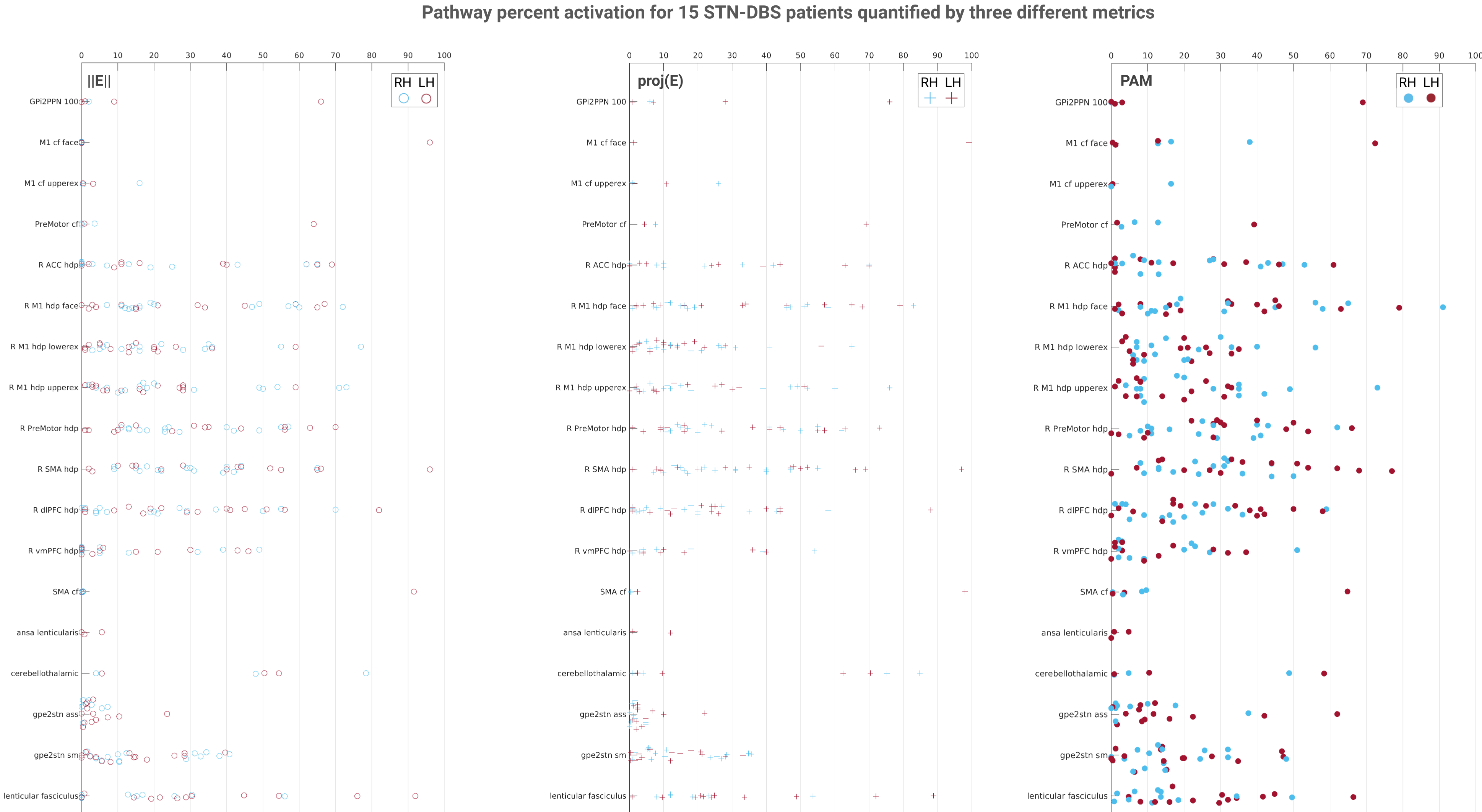

### Supplementary Figure S2

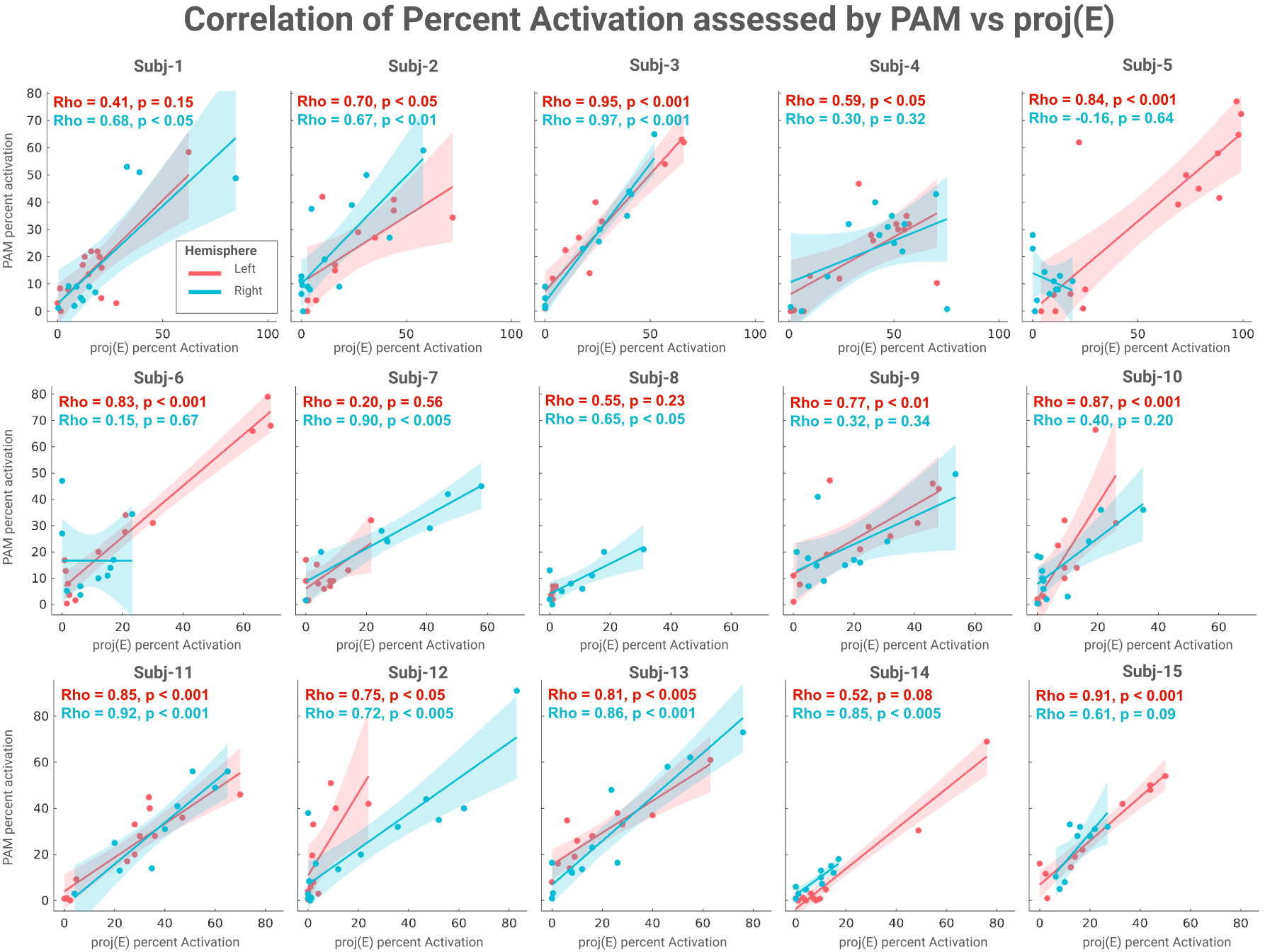

### Supplementary Figure S3

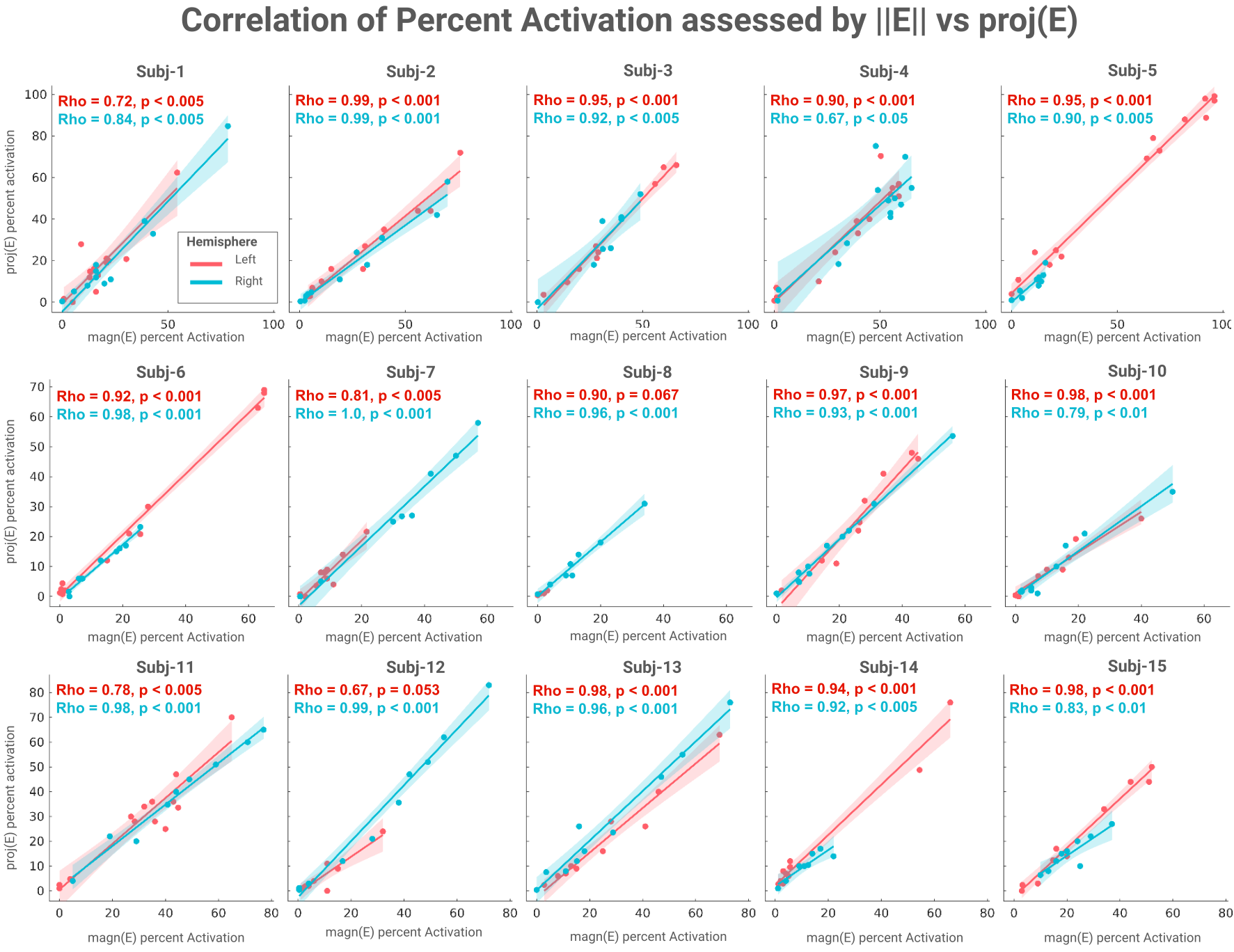
